# Comparing SARS-CoV-2 case rates between pupils, teachers and the general population: results from Germany

**DOI:** 10.1101/2021.03.04.21252877

**Authors:** Clemens Köstner, Stephan Letzel, Viktoria Eggert, Till Beutel, Pavel Dietz

## Abstract

Given the inconsistent state of research regarding the role of pupils and teachers during the SARS-CoV-2 pandemic in Germany, statewide and nationwide data of infection case rates were analyzed to contribute to the discourse. Infection data from official sources ranging from mid to late 2020 were collected, prepared and analyzed to answer the question if pupils, teachers and general population differed in active case rates or not. The data showed that pupils and teachers case rates didn’t exceeded those of the general population. In conclusion, it seems appropriate to appraise school-related measures to mitigate the SARS-CoV-2 pandemic sufficiently. Data-quality is a yet to overcome obstacle to provide good evidence-based recommendations regarding the management around infection cases in schools.

## Introduction

In March 2020, 1.5 billion pupils and their teachers worldwide were forced to stay away from schools as a result of measures to contain the spread of SARS-CoV-2.^1^ With the aim to positively influence the course of infection rates, schools in Germany were closed in March 2020 as response to the pandemic. This happened with far-reaching consequences not only for the education of the pupils, but for the entire society. Thenceforward, there was and still is a debate in Germany about the effectiveness of school closures and the role of children as drivers of the SARS-CoV-2 pandemic which led to several studies.

Some studies suggested that children and schools are not driving the pandemic and school-wide safety-measures (e.g., mask-wearing, distance) are sufficient to keep pandemic risks at an acceptably low level.^2–4^ Other studies showed that school closures are effective measures to slow down the pandemic.^5–7^ Based on this inconsistent state of research, the aim of the present study was to gather and analyze infection data from official sources to answer the question if pupils, teachers and the general population differ in active SARS-CoV-2 case rates.

## Methods

Official documentations of SARS-CoV-2 cases were analyzed. For the analysis, infection data provided by the Ministry of Education in Rhineland-Palatinate (BM-RLP), the Standing Conference of the Ministers of Education and Cultural Affairs (KMK) and the Robert Koch Institute (RKI), which is the government’s central scientific institution in the field of biomedicine in Germany, were used.^8–10^ Furthermore, some studies described that climatic conditions could have an impact on the spread of SARS-CoV-2, therefore this variable was included in the analysis.^4–5^ For this purpose, open access data of the daily average outside temperature from the federal German Weather Service (DWD) was used.^11^ The dataset for statewide (RLP) cases of pupils and teachers was started in August 2020, at the beginning of the school-year 2020/21 and ended in December, before the winter-break (data provided by the BM-RLP). The BM-RLP dataset for pupils and teachers was compared to the official dataset for infection cases of the Federal State Agency for Consumer & Health Protection of Rhineland-Palatinate (FSA-RLP).^12^ Cases in the BM-RLP dataset were interpreted as active cases since this was what schools were obliged to record. We calculated the estimate for daily active SARS-CoV-2 cases for the RLP general population by using the accumulated cases and subtracting the recovered cases (estimated by using 14 day as well as 23 day dropout algorithms, which are representing the lower boundary for the dropout algorithms used by the RKI and the lower boundary for the dropout algorithm used by the FSA-RLP) and the deceased cases. Further information regarding the methodology and a detailed list of limitations noticed during the analysis can be found in appendix 1.

The nationwide data for teachers and pupils was reported on a weekly basis and included five data-points of infection cases for a time period of five weeks starting in November and ending in mid-December 2020 (KMK).^9^ The KMK dataset was compared to the official dataset for infection cases in the general population of the RKI.^10^ We interpreted the nationwide weekly accumulated KMK SARS-CoV-2 cases for pupils and teachers as active cases and compared them to the respective active cases documented in the situation reports published by the RKI. To crosscheck the statewide findings for RLP, an analysis of the five available data-points for nationwide data of active cases for pupils and teachers from the KMK and general population from the RKI was performed.^9–10^

After identifying and obtaining the required SARS-CoV-2 data from official sources, school related data were transformed to bring all sources to a common denominator (i.e., rolling 7-day average estimate of active SARS-CoV-2 cases per 100,000). In the next step, the cases for pupils and teachers were subtracted from those of the general population for each analyzed day to generate distinct groups without autocorrelations. Then, state- and nationwide non-pharmaceutical interventions and school vacation periods were researched and incorporated into analysis and data presentation. Finally, information about the daily average outside temperature was integrated in order to take possible associations between the spread of the SARS-CoV-2 cases and outside temperature into account.

## Results

Figure 1 shows separate SARS-CoV-2 case rates for pupils, teachers and the general population, information about state- and nationwide non-pharmaceutical interventions, school vacations and the daily average outside temperature in the state of Rhineland-Palatinate (RLP).

**Figure 1:**
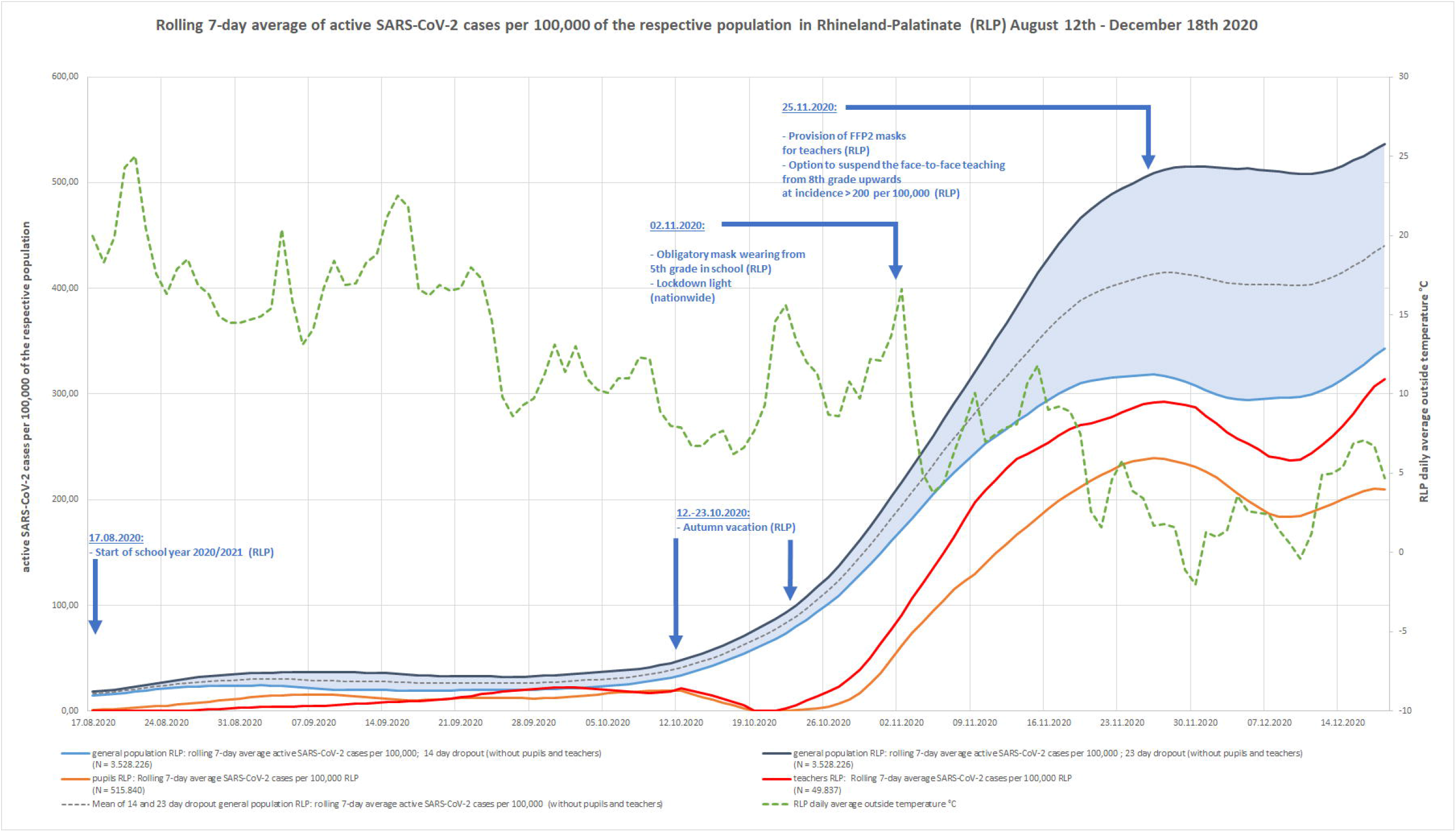
Rolling 7-day average SARS-CoV-2 cases per 100,000 of the respective population in Rhineland-Palatinate (RLP), August 12th - December 18th 2020

It can be seen that after relatively few SARS-CoV-2 cases in summer 2020, when schools reopened after the autumn vacation in late October 2020, active case rates in pupils and teachers moved below the estimated active case rates in the general population. In addition, there was an inverse proportional relation between the daily average outside temperature and estimated active cases of the general population, *r* = −.76, *p* = < .001 (14 day dropout estimation) and *r* = −.78, *p* = < .001 (23 day dropout estimation).

The crosscheck analysis for nationwide data provided the same relative positions of the three compared groups after subtracting pupils and teachers from the general population to create groups without autocorrelation. The general population showed the highest average active SARS-CoV-2 case rate per 100,000 people in that period of time (*M* = 389.19), followed by teachers (*M* = 351.35) and pupils (*M* = 192.03). Results for three bivariate correlations for the compared nationwide groups revealed no significant associations for the active SARS-CoV-2 cases between the three groups. On a non-significant level, pupils and teachers were more closely correlated, *r* = .78, *p* = < .12, than pupils and the general population, *r* = .14, *p* = < .82, and teachers and the general population, *r* = .21, *p* = < .74.

## Discussion

The results of our analysis imply that there were relatively more reported SARS-CoV-2 cases in the general population compared to pupils and teachers, especially after the autumn vacation. This pattern was found on a statewide and nationwide level of analysis. Knowing that the results for nationwide correlations between the compared groups are non-significant it seems reasonable to interpret those correlations in a way that pupils and teachers had to some extent more homogeneous infection case rates compared to the general population which showed a more independent course of case rates. Different explanations can be derived from the deviating rates of active SARS-CoV-2 cases of pupils and teachers relative to the general population. One might partly be the decrease in daily average outside temperature. This, for example, could have made the ventilation trough open windows in classrooms (which was mandatory for scools) more efficient.

The lower rates in pupils relative to teachers might be explained by an underestimation of cases among pupils due to the fact that SARS-CoV-2 infected children on average show fewer symptoms than adults or are even completely asymptomatic.^2^ This could have led to fewer testing of pupils compared to adults.

The ongoing narrative in the social discourse of pupils and teachers being relatively safe and not being drivers of the pandemic can be confirmed by our results. Nevertheless, recent studies targeting this topic showed that school closures slowed down the spread of SARS-CoV-2.^5–7^ In conclusion, it seems appropriate to continue to evaluate and improve school-related measures to mitigate the SARS-CoV-2 pandemic.

With regard to potential limitations, it would have been preferable to use higher quality data for state- and nationwide SARS-CoV-2 cases of pupils and teachers, as documentation procedures changed during the analyzed period. Furthermore, during the period of the autumn vacation, the documentation of SARS-CoV-2 cases for pupils and teachers was not continued, cases were artificially set to zero in the BM-RLP dataset. Another important limitation of our results is that the dropout algorithms for the subtraction of recovered SARS-CoV-2 cases from active cases used for the general population data (RKI & FSA-RLP) differ from the algorithm used for the school-based datasets (pupils and teachers). In the school-based datasets, active SARS-CoV-2 cases would drop out if the pupil or teacher continues to go to school (or uses a digital alternative), whereas for the general population dataset an algorithm estimates dropouts (e.g., RKI standard-dropout 14 days after a positive test, FSA-RLP standard-dropout 23 days after a positive test). Consequently, results should be interpreted with caution. A detailed and more in depth discussion of the potential limitations is given in appendix 1. It is important to consider, that SARS-CoV-2 cases of pupils and teachers do not necessarily indicate that the infections took place in schools.

Despite those limitations we conclude to further use the apparently useful school-related measures in order to mitigate the SARS-CoV-2 pandemic. In order to improve data quality, we encourage statewide officials to implement nationwide consistent methods of tracking and reporting SARS-CoV-2 infection cases of pupils, teachers and the general population to make them more adequately comparable. This would enable politicians in charge to better evidence-based decisions for the protection of pupils and teachers and to mitigate the SARS-CoV-2 pandemic in Germany.

## Data Availability

The BM-RLP data underlying this article were provided by Aufsichts- und Dienstleistungsdirektion (ADD) are open access and available online: https://add.rlp.de/de/corona-schulen/ueberblick-ueber-corona-infektionszahlen-an-schulen-in-rlp/. All other data (KMK & RKI) underlying this article are open access too and available via the URLs [9 & 10] shared in the references section

https://www.kmk.org/dokumentation-statistik/statistik/schulstatistik/schulstatistische-informationen-zur-covid-19-pandemie.html

https://www.rki.de/DE/Content/InfAZ/N/Neuartiges_Coronavirus/Daten/Fallzahlen_Daten.html;jsessionid=BFE70E3E9F65759B70884E18258973E0.internet102?nn=13490888

## Data availability

The BM-RLP data underlying this article were provided by Aufsichts- und Dienstleistungsdirektion (ADD). All data (BM-RLP, KMK, RKI, DWD & FSA) underlying this article are open access too and available via the URLs shared in the references section.^8–12^

## Key points

- Pupils and teachers showed lower SARS-CoV-2 case rates than the general population.
- The present paper provides a detailed list of potential pitfalls regarding the comparison of school related SARS-CoV-2 data with data of the general population.
- Data-quality is a yet to overcome obstacle to provide good evidence-based recommendations regarding the management around infection cases in schools.
- Apparently useful school related measures to mitigate the SARS-CoV-2 pandemic should be continued.

