## Appendix 1 for "Comparing SARS-CoV-2 case rates between pupils, teachers and the general population: results from Germany"

**Appendix 1: a detailed list and more in depth discussion of the potential limitations and pitfalls noticed while analyzing the data for the present paper**

1. Active SARS-CoV-2 cases were the unit for our between group comparisons. The formula to calculate active cases for the state of Rhineland-Palatinate (RLP) was: cumulated SARS-CoV-2 cases minus deceased cases, minus recovered cases. The part of the formula where estimations were [indispensable](https://www.dict.cc/?s=indispensable) was the number of recovered cases, since there were (and still are) two distinct algorithms used by official institutions to calculate these active cases from the total cases of general population in RLP.

To make this point clear: the RKI calculated the number of recovered cases with a dropout algorithm starting to remove cases a) 14 days after entering the dataset if mild symptoms were reported, b) 4 weeks for pneumonia, and c) the end of hospitalization if hospitalization was reported. Since we had no further information available regarding the cases reported for the RKI dataset, a 14 day dropout was our best approximation for the estimation of active cases.

Federal State Agency for Consumer & Health Protection of Rhineland-Palatinate (FSA-RLP) used a dropout algorithm starting to remove recovered cases with a different definition of steps and different dropout-delays. Recovered cases were defined as follows: a) non-deceased, non-hospitalized, and ill more than 21 days ago from data cutoff date; b) non-deceased, reported hospitalized and ill more than 28 days ago from data cut-off date; c) non-deceased, hospitalization unknown and ill more than 28 days ago from data cut-off date. If no date of illness was available, the estimated date of illness from reporting date minus a mean reporting delay of 5 days was used. Since there was no information available regarding hospitalization, we used c) as the standard case for the FSA-RLP dataset (28 days). In the next step we corrected those 28 days by subtracting 5 days, because there was no data on the date of illness available. So for the FSA -RLP dataset, a 23 day dropout was our best approximation for estimating active cases.

From the circumstances described above, it is obvious that estimates of active cases for RLP differ between the RKI and FSA-RLP. Even today, in September 2021, there are still deviating officially reported numbers of active cases for RLP from the RKI and the FSA-RLP.

Since there was no further information available to specify the category (e.g., pneumonia: yes /no) and date of illness for official records of SARS-CoV-2 cases, we used two variations of dropout algorithms as lower and upper boundaries (i.e., 14 days and 23 days) for our estimations. To be transparent regarding this issue, we plotted both, curves and their mean for active cases in RLP in Figure 1. We do think that since the values for recovered cases (which are the basis for active cases) are estimates, it is reasonable to assume that the range between the estimates of the RKI and the FSA-RLP algorithms is most likely a good [approximation](https://www.dict.cc/?s=approximation) of the active SARS-CoV-2 cases.

To validate our assumptions empirically, we compared our (as described above) calculated mean estimates for active cases in RLP for August 2021 and the official data provided by the FSA-RLP for active cases (unfortunately the available documentation for active cases in RLP has just started in August 2021). Our estimations and the reported official data for August 2021 only had a 5.1% deviation. We do think that given the fact that some assumptions had to be made to calculate the active cases estimates a 5.1% deviation was acceptably adequate and represents the best approximation we could achieve.

2. The mentioned differences in dropout calculations from the RKI and the FSA-RLP were not the only difficulty by estimating the active cases. The dataset for pupils and teachers provided by the BM-RLP used an “empirically dropout” , i.e., the recording of a case started when a pupil or teacher was [absent](https://www.dict.cc/?s=absent) [due](https://www.dict.cc/?s=due) to [illness](https://www.dict.cc/?s=illness) and ended when the person was back at school. As one can easily see, there was no perfect match of the different datasets in use.
3. Another aspect regarding the datasets that is not easily comparable: entering the datasets. Schools in RLP had the obligation to record all active SARS-CoV-2 cases in pupils and teachers that led to absence from school. For schools it was not part of the standard procedure to check or verify, e.g., via documented PCR test, that one had a SARS-CoV-2 infection. We do think that it is reasonable to assume that most of the pupils and teachers that claimed to have a SARS-CoV-2 infection really had it and that most of the cases with SARS-CoV-2 reported this to the school, but there is certainly room for doubt.

On the other side the process to enter the RKI or FSA-RLP datasets was different: a laboratory confirmed positive PCR test was necessary. Laboratories reported positive SARS‑CoV‑2 cases directly to the local public health departments which then reported the numbers to the RKI.

Given that knowledge, comparisons between school-related and general population-related datasets become less convincing.

4. Another important limitation to our study is the switch in the process of documenting SARS-CoV-2 cases for pupils and teachers in RLP. Before the autumn vacation in RLP officials from the Ministry of Education documented cases for all schools in RLP on the basis of information they could get from schools. After the autumn vacation schools themselves where obliged to document and report the daily number (without recording Name or unique ID) of all known active SARS-CoV-2 cases of pupils and teachers to the Ministry of Education RLP (except for weekends where documentation stopped / autumn vacation where all documented case-numbers were artificially set to zero) by using an online-tool. Therefore the timespan before and after the autumn vacation is not based on a consistent process for documenting SARS-CoV-2 cases for pupils and teachers in RLP.
5. Even though it is obvious that all the above mentioned limitations had an impact on the calculated results reported in our paper, there was a lack of better alternative dataset to choose from. We gave our best to be as precise and transparent as possible when analyzing the data and writing the manuscript and the appendix. Furthermore, we used the – to our knowledge – best possible way to estimate active cases from heterogeneous data-sources, knowing that with more consistent data our estimations would have been better or even unnecessary (if all infections would have been recorded und reported equally).
